# 31P-MRS of healthy human brain: revealing the hidden PME signals under phosphoethanolamine and phosphocholine resonances at 7T

**DOI:** 10.1101/2022.06.19.22276613

**Authors:** Jimin Ren

## Abstract

**Purpose:** For decades, it has been common practice to quantify brain phosphomonoester (PME) 31P signals by a two-component model composed of phosphoethanoamine (PE) and phosphcholine (PC). This study presented the evidence of hidden PME (h-PME) signals and their selective detections toward accurate quantification of PE and PC.

**Methods:** Gaussian lineshape analysis was modeled to reveal h-PME. Inversion-recovery (IR) sequence was employed to null the PE and PC resonances for selective detection of h-PME. The fully-relaxed 31P spectra after h-PME correction were used to quantify PE, PC and other brain metabolites in a group of 16 healthy subjects.

**Results:** Spectral lineshape analysis and IR modulation revealed previously overlooked h-PME signals underlying sharp PE and PC resonances. The h-PME signals appeared as a broad “bump” (LW1/2: 105 ± 25 Hz, N = 16), leading to poor spectral resolution between PE and PC. Fast relaxing h-PME signals, tentatively assigned to blood 2,3-DPG, were selectively detectable using IR sequence at an optimal inversion delay of 5.8 s. In fully relaxed 31P spectra, h-PME measured 44 ± 9 % of the total PME signal, equivalent to 1.36 ± 0.39 mM in single phosphoryl unit, compared to 1.41 ± 0.23 mM for PE and 0.31± 0.10 mM for PC.

**Conclusion:** Hidden PME signals are a significant constitute of PME signals in human brain 31P spectra, and need to be taken into account for quantifying PE and PC as biomarkers of altered phospholipid metabolism in brain pathologies.

## 1. Introduction

Phosphoethanolamine (PE) and phosphocholine (PC) are essential precursors and degradation products of phosphotidylethanolamine (PtE) and phosphotidycholine (PtC) – the major constituents of membrane phospholipids (MPL, Figure 1).^1^ PE and PC are also important in cellular neurochemistry by interactions with blood-brain-barrier permeable substrates and cellular signaling molecules (such as FFA, PA and DAG).^2^ Alterations of brain MPL metabolism and neurochemistry have been associated with brain development,^3^ cancer,^4, 5^ and a number of neurological disorders such as Alzheimer’s disease,^6, 7^ Parkinson’s diseases,^8, 9^ schizophrenia,^10, 11^ and multiple sclerosis.^12, 13^

**Figure 1.**
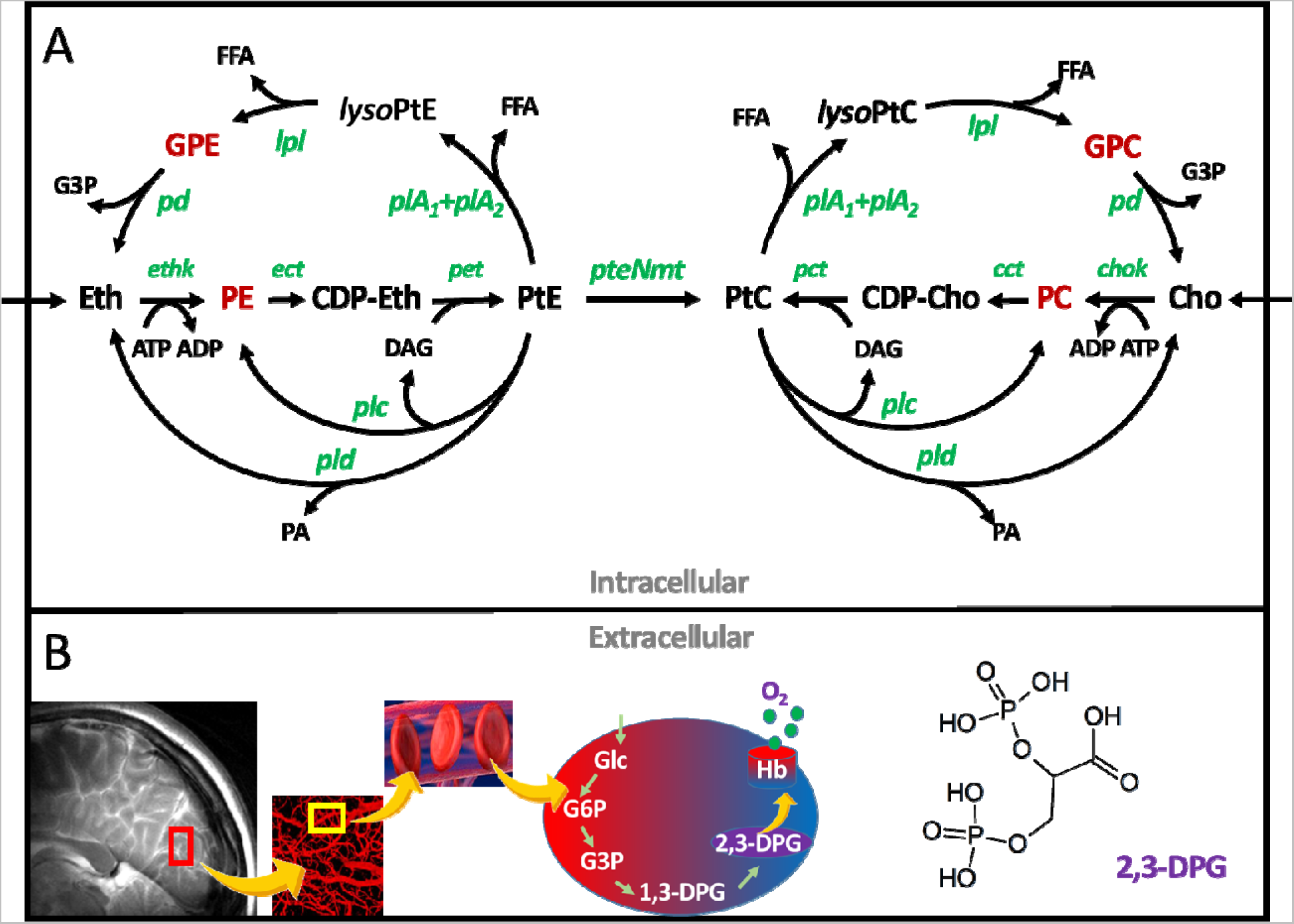
(A) Schematic of phospholipid metabolic pathways involving PE and PC and their counterparts GPE and GPC. (B) Diagram showing the physiological basis for the observation of 2,3-DPG signals in human brain. Abbreviation: 2,3-DPG, 2,3-bisphosphoglycerate; Cho, choline; Eth, ethanolamine; PC, Phosphocholine; PE, phosphoethanolamine; CDP-Cho, cytidylphosphone-choline; CDP-Eth, cytidylphosphoethanolamine; FFA, free fatty acid; G3P, glycerol 3-phosphate; GPC, glycerolphosphocholine; GPE, glycerolphosphoethanolamine; lysoPtC, lysophosphotidylcholine; lysoPtE, lysophosphotidylethanolamine; PA, phosphotidic acid; MPL, membrane phospholipids. Enzyme abbreviation: chok, choline kinase; ethk, ethanolamine kinase; cct, choline-phosphate cytidyltransferase; ect, ethanolamine-phosphate cytidyltransferase; cpt, choline-phosphotransferase; ept, ethanolamine-phosphotransferase; lpl, lysophospholipase; pd, phospholipase D; plA1, phospholipase A1; plA2, phospholipase A2; and pteNmt, phosphotidylethanolamine N-methyltransferases.

In vivo 31P-MRS is currently the only imaging modality capable of simultaneously measuring PE, PC, together with glycerophosphoethanolamine (GPE), phosphocholine (GPC), PCr and ATP, thus offering an integrated view of brain MPL metabolism neuroenergetics.^14^ For decades, the 31P-MRS method for quantifying PE and PC has been based on a two-component model, assuming that the 31P signals in phosphomonoester (PME) region are composed of only PE and PC, similar to the counterpart phosphodiesters (PDE) region composed of only GPE and GPC. This understanding has been cemented as prior knowledge in brain 31P spectral analysis based on LCModel and AMREA – two widely used quantitation tools.^15, 16^ However, there has been a growing body of 31P data showing spectral complexity in PME region in contrast to PDE, especially at ultrahigh fields.^17–25^ Typically, PE and PC are poorly resolved, whereas GPE and GPC, at approximately equal peak separation (0.5 ppm), are well-resolved. Besides, measurement discrepancies have been noticed for metabolites in PME region.^16, 19, 23, 26^ Moreover, additional signals distinct from PE and PC have been documented in 31P NMR studies of brain tissue extracts.^27–30^ All these raise the question whether there are really hidden PME (h-PME) signals in brain 31P spectra in vivo.

The present study aimed to reveal h-PME signals under PE and PC in brain 31P spectra in vivo, using lineshape analysis and inversion-recovery method. Concerning the sources of h-PME signals in vivo, a likely contributor is blood 2,3-DPG, as implied by recent studies showing apparent T2 shortening at PC and signal attenuation at PE upon OVS suppression of periphery tissues with large vascular spaces.^24, 26, 31^ Over the last three decades, a few attempts have been made to include 2,3-DPG in PME fitting,^32–34^ though they differ widely on the details of lineshape model, due to a lack of obvious spectral features assignable to 2,3-DPG. In this study, the strategy to selectively detect h-PME was based on the T1 shortening of 2,3-DPG in RBC,^35, 36^ rather than on the classic resonance pattern of 2,3-DPG, considering that the resonance frequencies of 2,3-DPG in RBC may be dispersed due to sensitivity to variations in pH, oxygenation, and magnetic anisotropic susceptibility effect from spatial orientation of blood vessels.^24, 31, 35–38^ The in vivo brain spectral features documented in this study clearly demonstrated complexity of PME composition and inadequacy of the conventional two-component model. The finding of h-PME may have implications in using PE and PC measurements as biomarkers of altered MPL metabolism in neuropathology.

## 2. Materials and Methods

### 2.1 Protocol approvals and consent

This study was approved by the Institutional Review Board of The University of Texas Southwestern Medical Center. Prior to the MRS study, informed written consent was obtained from all participants. Sixteen participants (nine male and seven female), aged 38.5 ± 12.0 years, BMI 28.4 ± 4.3 kg/m^2^, resting heart rate 73 ± 14, and peripheral capillary oxygen saturation (SpO2) 96 ± 3% participated in the study. All subjects were in good general health with no history of peripheryl vascular, systemic, myopathic, cancer, psychiatric, or neurodegenerative diseases. Heart rate and blood oxygen saturation level were monitored during the scan.

### 2.2 Protocol and data acquisition

All subjects were positioned head-first and supine in the human MRI scanner (7T Achieva, Philips Healthcare, Best, the Netherlands), with the back of the head positioned in the center of the detection RF coil (Philips Healthcare). The RF coil was a partial volume, double-tuned 1H/31P quadrature T/R coil consisting of two tilted, partially overlapping 10-cm loops. Axial, coronal, and sagittal T2w turbo spin echo images were acquired for planning voxel shimming. Typical MRI parameters include field of view (FOV) 180 x 180 mm^2^, repetition time (TR) 2.5 s, echo time (TE) 80 ms, turbo factor 15, in-plane spatial resolution 0.6 x 0.7 mm^2^, slice thickness 6 mm, number of acquisition (NA) one, and acquisition time 2.1 min. Second order 1H-based automatic volume shimming was applied prior to 31P spectral acquisitions.

The time-domain free-induction-decay (FID) ^31^P data were acquired using a block-shaped excitation pulse with B1max = 59 μT, pulse width pw = 22 μs, flip angle θ = 55°, excitation bandwidth = 6.5 kHz, receiver bandwidth = 8 KHz, TR = 30, and NA = 16. The dead-time (DT) of the readout pulse prior-to FID sampling was minimal (0.17 ms) or set to 0.5-ms to avoid background MPL background signals. The effect of TR on PME signals were compared between TR = 1 s and 15 s, with NA = 256 and 16 s, respectively. Localized 31P spectra were acquired using 2D MRSI sequence with TR = 2 s, slice data matrix 15 x 15, FOV 18 x 18 cm^2^, TE = 0.5 ms, and eight scan averages. Three additional outer-volume-saturation bands were applied to suppress possible signal contamination from extra-cranial tissues and blood circulating in the sinuses of the brain posterior. An inversion-recovery (IR) experiment was carried out for selective observation of h-PME signals. The IR sequence was composed of a 5-ms trapezoid-shaped adiabatic pulse with inversion bandwidth of 7 kHz, followed by a varying delay period TI, and a hard readout pulse. The inversion delay TI was optimized to null MPL metabolites for profiling h-PME.

### 2.3 31P Spectral analysis

The frequency-domain spectra were obtained by Fourier transformation (FT) of FID data following apodization, zero-filling of sampling points (from 4k to 8k), and phasing (zero-and first-order). For data acquired at minimal DT, the first three FID data points were discarded prior to FT to remove broad MPL background signals. After post-processed using the scanner software SpecView, the 31P spectra were further analyzed using an in-house program written in Matlab (2021b, The MathWorks, Inc. Natick, MA), which is featured with baseline correction and lineshape fitting. Gaussian lineshape model was used for fitting all 31P signals, each defined by three parameters including chemical shift δ, linewidth LW_1/2_, and height h. The α-, β- and γ-ATP signals were fitted by doublet (α- and γ-) or triplet with a fixed J(31P-31P) coupling constant of 17 Hz. h-PME signals were modelled using two Gaussian lineshapes. Baseline with broad background signals was determined by interpolation of pivotal points selected outside targeted signals using a combination of *spline* and *pchip* functions. The fitting parameters derived from the group-summed 31P spectrum were used as initial fitting parameters for analysis of spectra acquired from individual subjects (N = 16). To obtain spectra with fully resolved PE and PC signals, spectra acquired with the IR sequence optimized for detection selective of h-PME were subtracted from reference spectra acquired without inversion. Peak integrals (area-under-curves) derived from fully relaxed spectra at TR = 30 s were used to calculate metabolite concentrations, in reference to γ-ATP as an internal standard (3.0 mM). PCr resonance was used as the chemical shift reference (δ = 0 ppm).

### 2.4 Statistical analysis

All data were reported as mean ± standard deviation, calculated using MATLAB.

## Data Availability Statement

The data are available from the corresponding author upon reasonable request.

## 3. Results

### 3.1 Revealing h-PME by lineshape analysis

#### 3.1.1 Non-localized spectrum at short TR

In vivo brain 31P spectra clearly demonstrated a striking contrast in spectral resolution between MPE and PDE regions, as shown in Figure 3 for the case of non-localized data acquired at short TR (1s). while GPE (3.50 ppm) and GPC (2.95 ppm) in PDE region were fully resolved and fit well by Gaussian linehshape with two-component model (LW1/2: 16.8 and 16.0 Hz), their counterparts PE and PC in PME region (6.76 and 6.21 ppm), at comparable peak separation (∼0.5 ppm), were resolved only partially (76% and 54%) and deviated considerably from two-component Gaussian model. However, with inclusion of two additional Gaussian component ((δ: 6.83 and 6.43 ppm; LW1/2: 53.6 and 62.1 Hz; integral 1:1), the PME fitting performed equally well as PDE, yielding PE and PC linewidths (LW1/2: 16.9 and 16.1 Hz) nearly identical to those of GPE and GPC and with small and evenly distributed residuals (Figure 3(B)). The sum of these two additional PME signals appeared as a broad “bump”, accounting for 59% of total PME signals in this particular case. In comparison, PE and PC were 31% and 10% of total PME, respectively.

**Figure 2.**
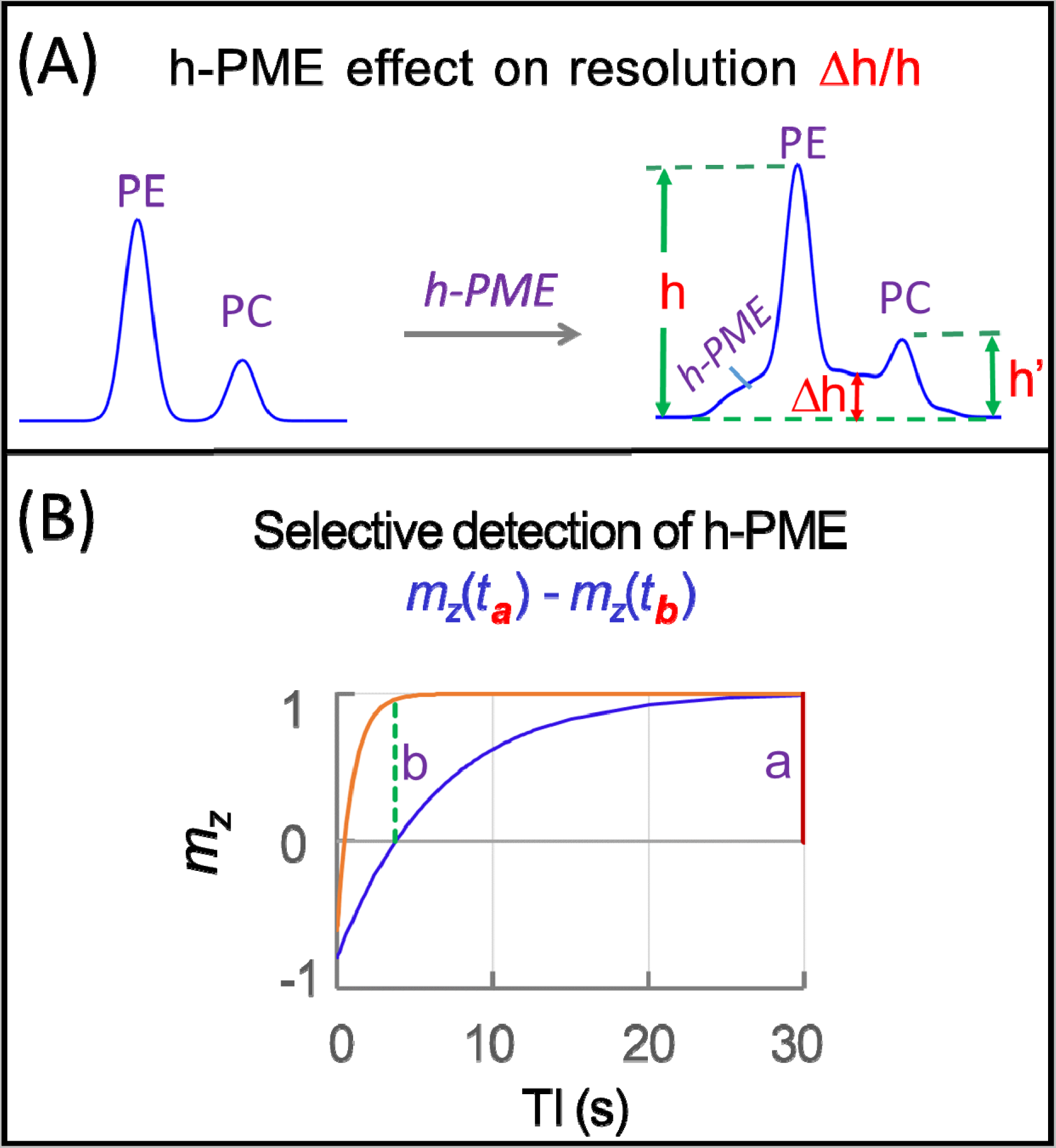
(A) Illustration of the definition of apparent spectral resolution at PE and PC. (B) Schematic representation of the basic principle of selective detection of fast-relaxing hidden PME (h-PME) signals by inversion-recovery modulation.

**Figure 3.**
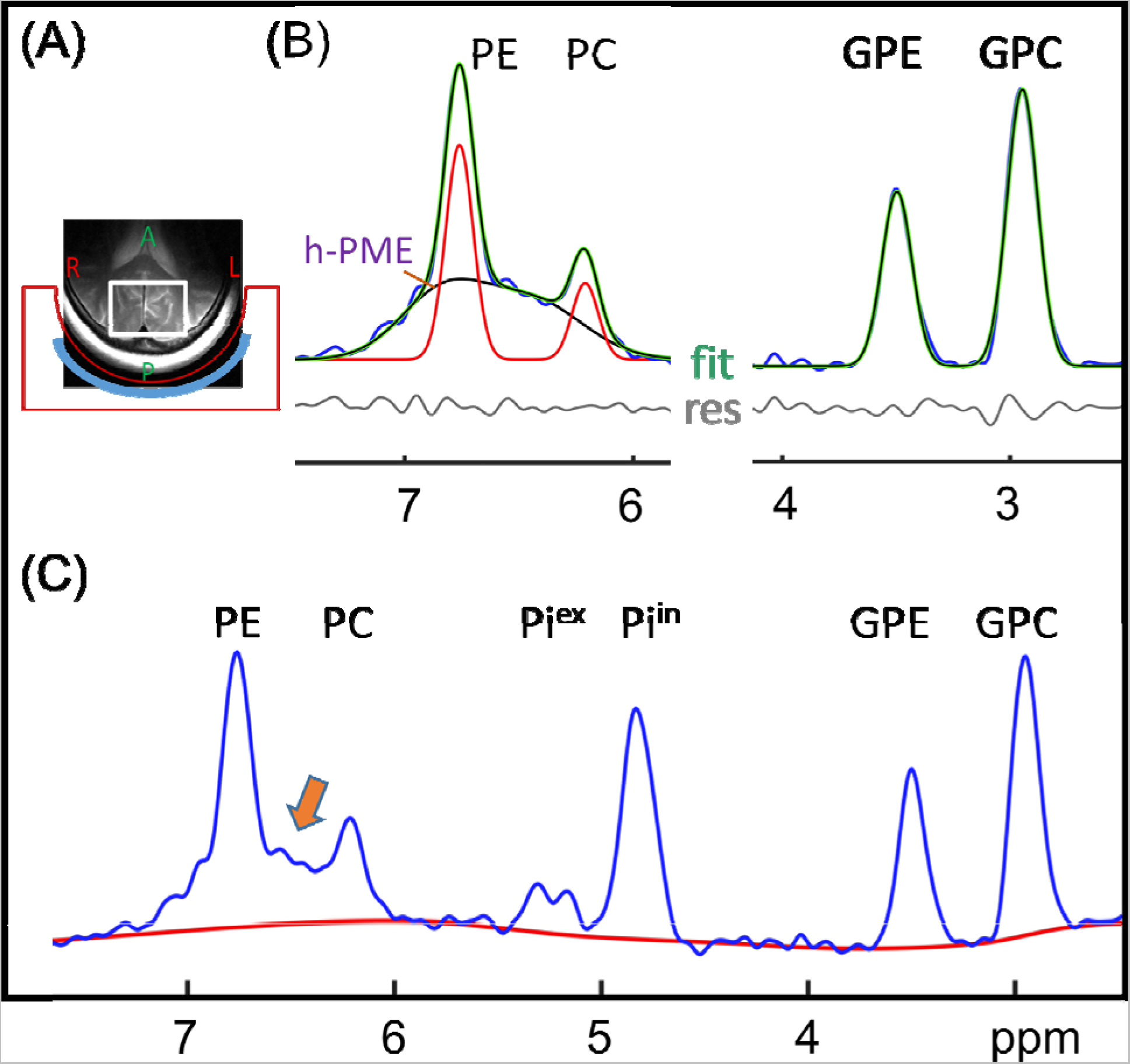
Lineshape analysis of PME and PDE signals. (A) Illustration of RF coil setting for acquiring brain 31P MR spectra, with an axial T2w image showing the placement of shimming box (white square). (B) Gaussian lineshape analysis of PME (left) and PDE (right) signals. Note the contrast between PME and PDE regions in Gaussian lineshape fitting. A broad h-PME “bump” is seen in PME region as residuals of two-component model fitting. (C) Regional 31P spectrum with baseline added to reveal the poorly resolved PE and PC signals in contrast to well-resolved GPE anf GPC signals. Data collected from a 23 years old man with a pulse-acquire sequence at short TR of 1 sec and 512 scan averages.

#### 3.1.2 Localized spectrum at short TR

A similar resolution contrast was also observed between PME and PDE in data acquired by localized 2D 31P MRSI, as shown in Figure 4 where the peak resolution measured 81% at PE and 69% at PC, whereas GPE and GPC were fully resolved. In this particular case, the lineshape fitting with inclusion of two additional PME signals (1:1 in integral) yielded equal linewidth (17.6 Hz) for PE and PC, comparable to GPE (19.3 Hz) and GPC (19.0 Hz) in PDE region. Again, the sum of two hidden PME components (48.0 Hz at δ = 6.88 ppm, and 65.4 Hz at 6.46 ppm) formed a broad “bump”, making up 47% of total PME, compared to PE 39% and PC 14%.

**Figure 4.**
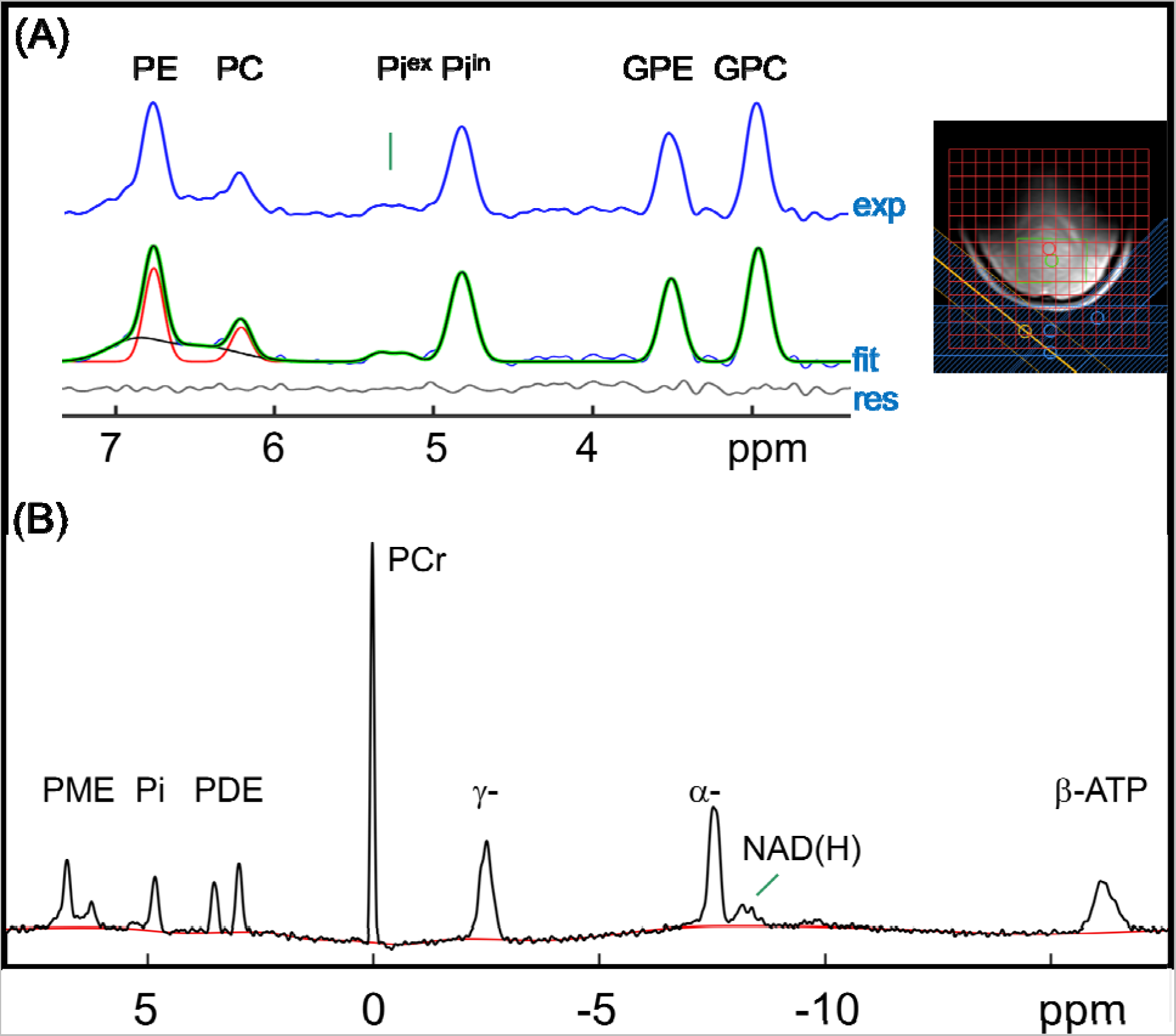
Lineshape analysis of 31P spectra acquired by 2D MRSI. (A) A voxel-summed spectrum and the lineshape fitting by Gaussian model. Note a broad “bump” is seen underlying PE and PC as fitting residuals of two-component Gaussian model, whereas GPE and GPC fit well to the same two-component model. The inset shows the layout of the 2D MRSI data matrix, the placement of OVS bands (in blue), and the location of selected voxels (white box). (B) Full 31P spectrum with baseline added (red trace) to highlight the resolution contrast between PME and PDE regions. Data from a summation of 20 voxels centered in the occipital-parietal region, acquired by 2D MRSI at TR = 2 s.

#### 3.1.3 Short vs long TR non-localized spectra

An increase in TR tends to decrease the h-PME proportion of total PME and consequently improve resolution between PE and PC, as shown in Figure 5. In this case, the hidden PME signals measured 47% of total PME at TR 15 s, lower than 62% measured at TR 1s. In comparison, PE and PC were 44.5% and 8.6% of total PME at TR = 15 s, respectively, reduced to 31% and 7% at TR = 1 s, respectively. The corresponding apparent peak resolution was 83% for PE and 60% for PC at TR = 15 s, compared to 72% for PE and 45% for PC at TR = 1 s.

**Figure 5.**
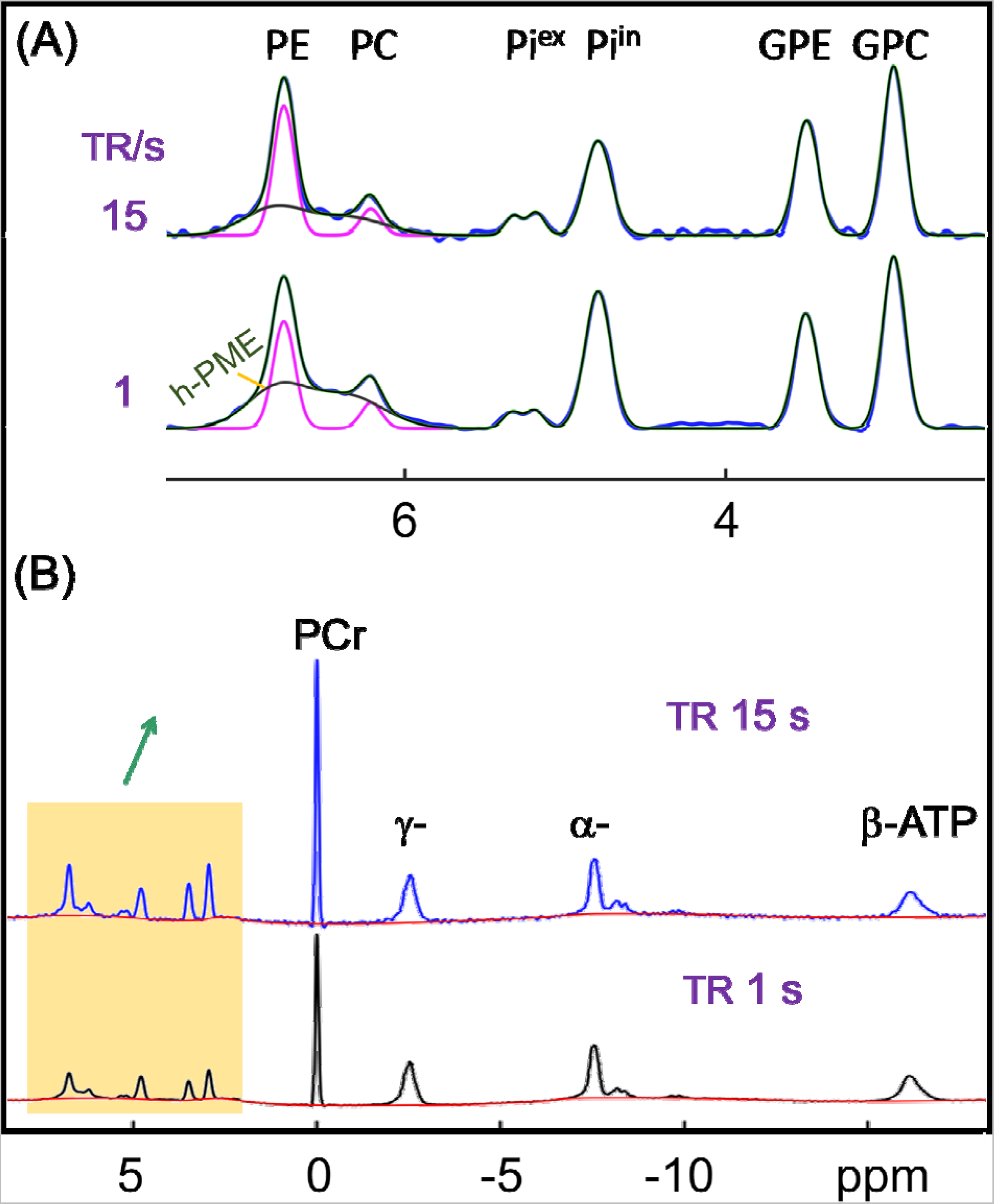
TR effect on h-PME and spectral resolution between PE and PC. (A) Downfield 31P signals showing decrease in h-PME proportion and improvement in PE and PC resolution as TR increase from 1s to 15 s. (B and C) Screenshots of full-range 31P spectra acquired at TR of 15 s (B) Full-range 31P spectra with baseline shown in red trace. The data were acquired from a 58 years old male with NA = 16 (B) and 512 (C).

### 3.2 Selective detection of h-PME

The fact that h-PME becomes less populated at longer TR indicates that h-PME has short T1 compared to PE and PC, and this feature was exploited to selectively detect h-PME by IR modulation. As shown in Figure 6(A), compared to the reference spectrum without inversion, the IR spectrum acquired at an inversion delay of 5.8 s was distinctly different in the downfield region, especially the PME region which was featured with a broad “bump” from h-PME, with PE and PC nearly completely nulled. In contrast, there was no change in the GPE-to-GPC ratio, except the scaling factor. In addition to h-PME, all the upfield short-T1 signals were fully recovered including α-, β-, and γ-ATP and NAD(H) all fully recovered (Figure 6(B)). The dynamic changes of PME and PDE signals in response to inversion delay are shown in Figure 1S (Supplementary Material).

**Figure 6.**
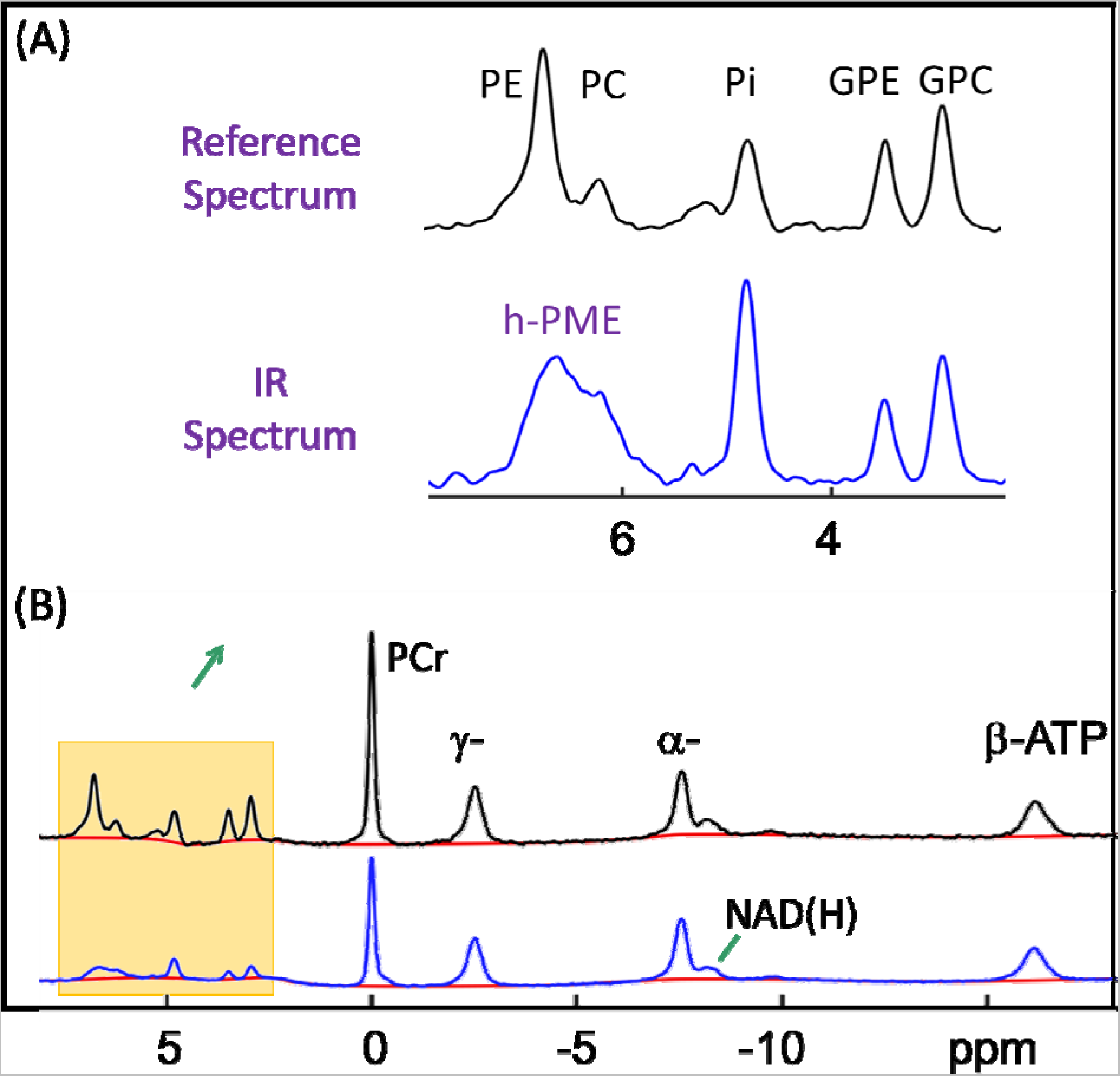
Selective detection of fast-relaxing h-PME. (A) Comparison of reference spectrum without inversion (top, black trace) and inversion-recovery (IR) spectrum with nulled PE and PC (bottom, blue trace, vertically scaled on magnitude of PC for easy comparison). (B) Full-range reference (top) and IR (bottom) 31P spectra. The data were acquired at long TR of 30 s, summed over 6 subjects. An inversion delay of 5.8 s was used for IR data acquisition. Note the full recovery of ATP and NAD(H) signals in contrast to slow recovery of MPL metabolites (PE, PC, GPE and GPC).

### 3.4 Filtering out h-PME to resolve PE and PC

To filter our h-PME, a subtraction was performed between the reference spectrum and IR spectrum. As shown in Figure 7, the subtraction led to fully resolved PE and PC after baseline correction and the removal of residual signals. Interestingly, in addition to spectral resolution, the other outcomes of this spectral editing were also not critically sensitive to the inversion delay, as compared in Figure 7(A) (TI = 5.8 s) and 7(B) (TI = 3.8 s). For example, the measured linewidths (LW1/2) at TI = 3.8 s were 18.3, 18.0, 20.3 and 19.4 Hz for PE, PC, GPE and GPC, respectively, comparable to 18.2, 17.9, 20.1 and 19.3 Hz at TI = 5.8 s. For different MPL metabolites, the relative signal intensities were PE : PC : GPE : GPC = 1.33 : 0.25 : 0.80 : 1:00 at TI = 3.8 s, vs 1.42 : 0.27 : 0.77 : 1.00 at TI = 5.8 s.

**Figure 7.**
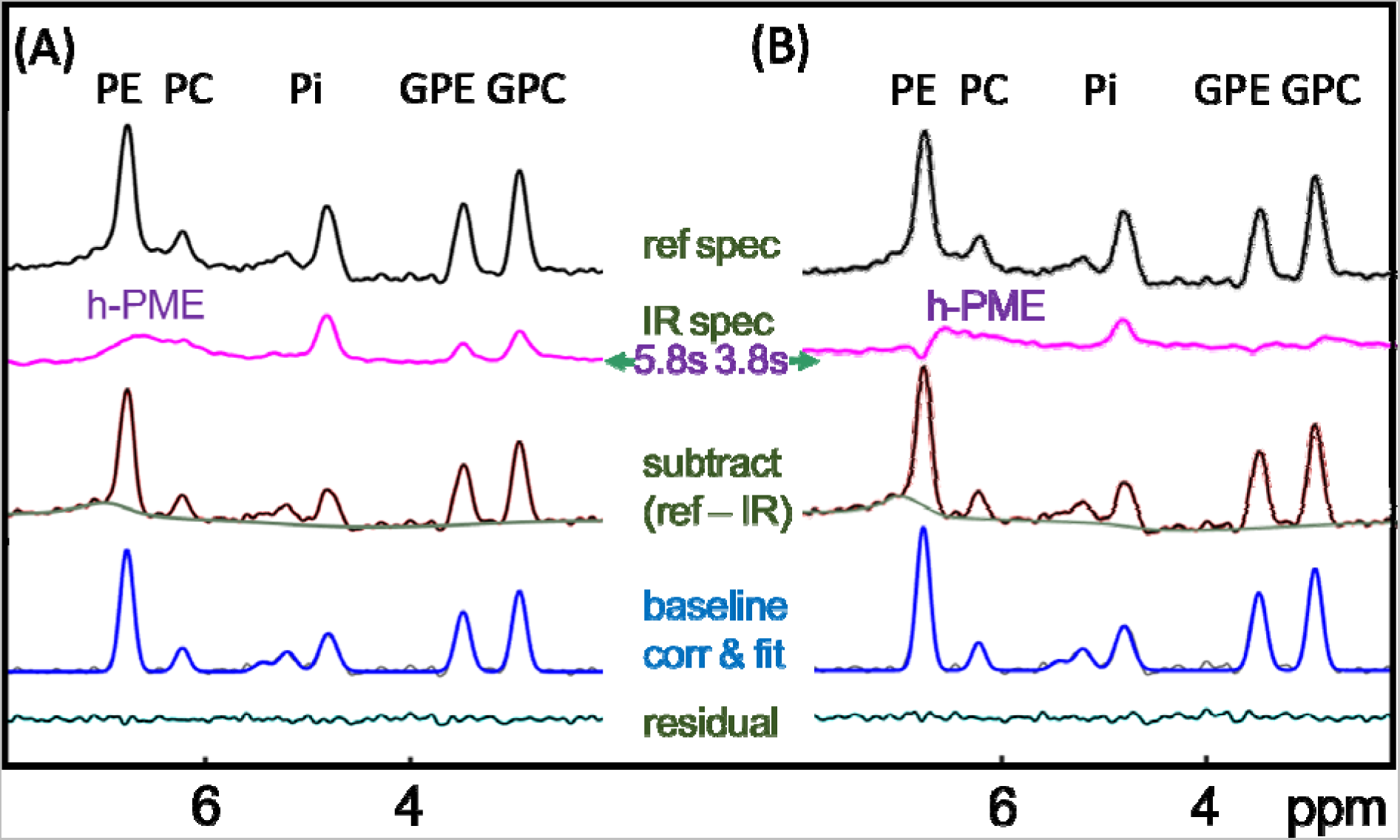
Effect of inversion delay time (TI) on PME recovery, peak resolution and lineshape for TI = 5.8 s (A, left panel) and 3.8 s (B, right panel). Note that PE and PC are fully resolved in subtraction spectra for both TIs and that there is a residual “bump” in the baseline, representing slow-relaxing h-PME component. The summation of the “bump”-containing baseline and the IR spectra are shown in Figure 8.

### 3.5 Profiling the entire h-PME

In addition to a broad, fast relaxing h-PME component observed in the IR spectra, there was another minor but slowly relaxing component in the subtraction spectra, visible as a baseline background signal at the foot of PE peak (Figure 7). A combination of these two h-PME components gave rise to the profile of the entire h-PME, as illustrated in Figure 8(A). As expected, regardless of the dataset (TI = 5.8 s vs 3.8 s) for the combination, the resultant h-PME profiles were very similar in size and linewidth (average LW1/2 = 108.5 Hz, Figure 8(A) and (B)). It was found that that maximum of the h-PME profile was coincident with PE resonance. Interestingly, in addition to h-PME, the same datasets also reveal a fast relaxing h-PDE signal with similar linewidth (average LW1/2 = 110.6 Hz) centered at 2.2 ppm, likely from mobile phospholipids or phosphoproteins as noted in literature.

**Figure 8.**
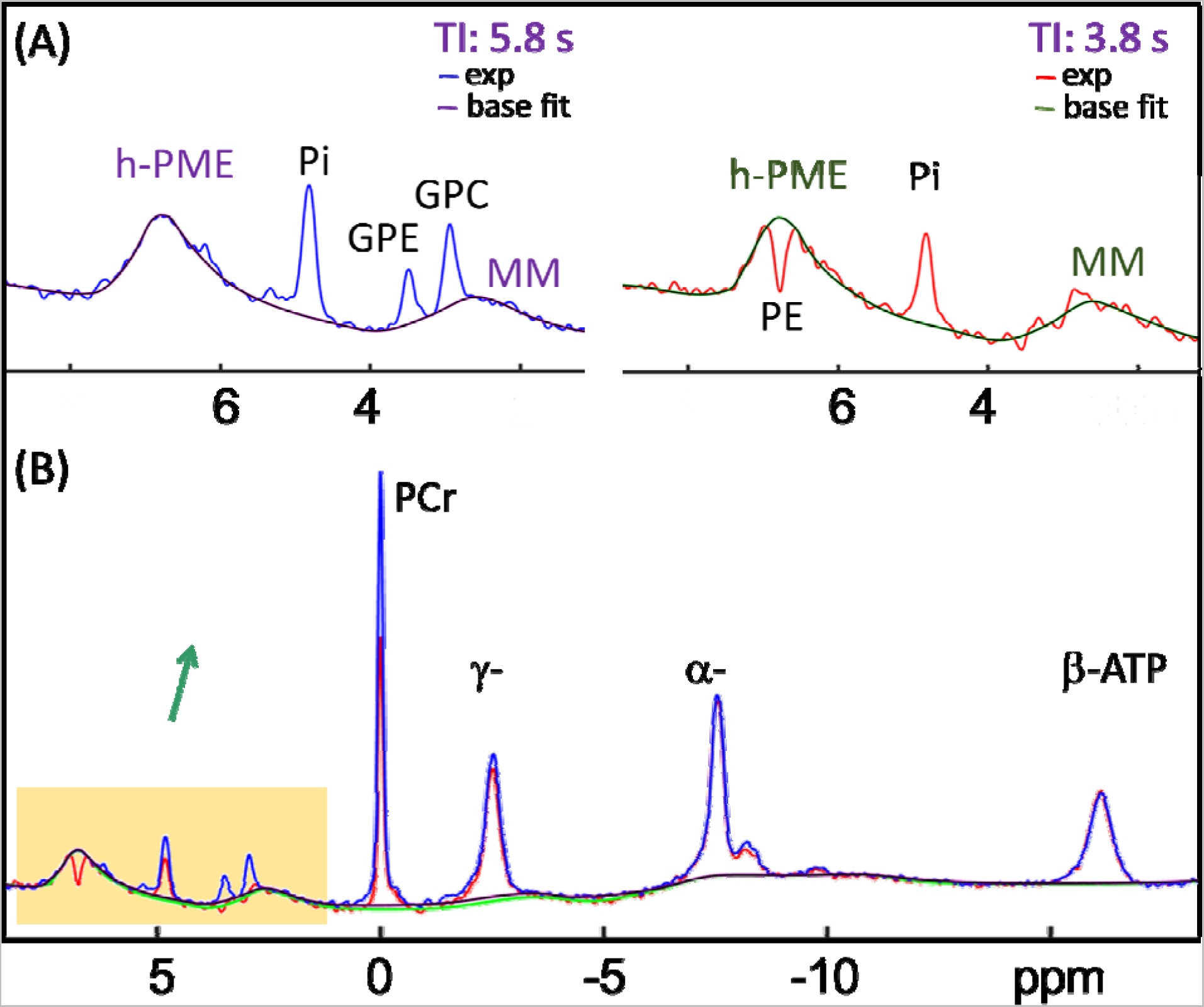
Inversion-recovery spectra summed with baseline signals for profiling h-PME. (A) Profiling h-PME and an unspecified macromolecule signal (MM) in the upfield of GPC for datasets TI = 5.8 s (left panel) and 3.8 s (right panel). (B) Full-range spectra with two datasets overlaid to show nearly identical h-PME profile and full recovery of ATP signals.

### 3.5 h-PME correction for quantification of PE and PC

The obtained h-PME profile was used to correct h-PME contribution in spectra acquired without inversion but under otherwise the same sequence condition, taking into consideration of scaling factors. This led to fully resolved PE and PC in PME region that is convenient for quantification and comparison with other metabolites, as shown in Figure 9 for a group of subjects (28 scans from N = 16 subjects). The averaged h-PME was 43.6 ± 8.8 % of total PME under fully relaxed condition (TR = 30 s). The combined concentration of PE and PC was 1.72 ± 0.29 mM, as compared to 3.07 ± 0.45 without correction (total PME). Table 1 summarizes the results of quantitative analysis for the entire group, and compared with those in literature without correction, in reference to γ-ATP (3 mM). Supplementary Material Table 1S summarizes the measured linewidth (LW1/2, in Hz) for all observed metabolites.

**Figure 9.**
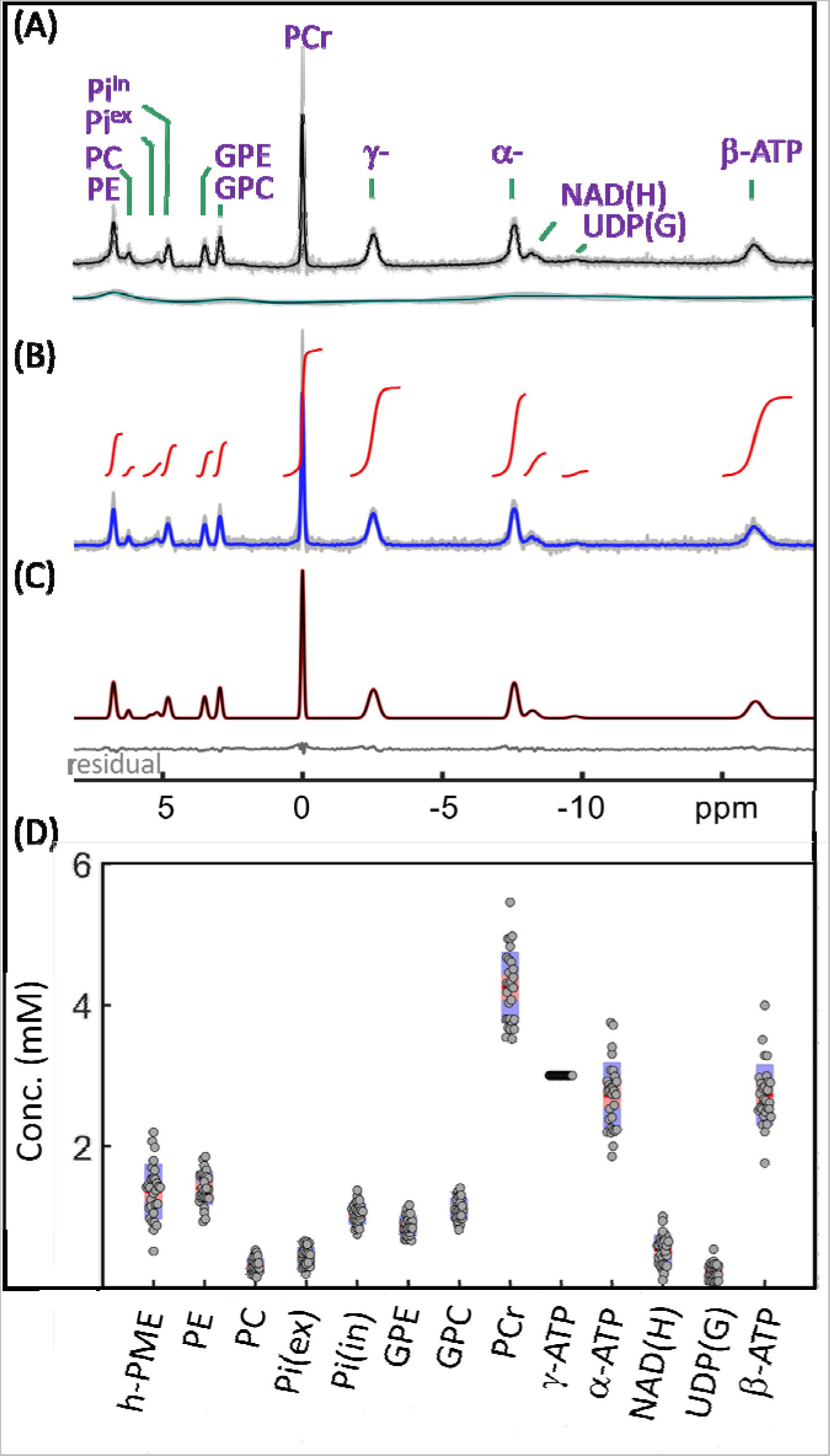
(A) Brain 31P MR spectra prior-to (A) and post (B) correction of h-PME and other background signals for 28 scans from N = 16 subjects. Data acquired at TR = 30 s and scaled on integral of γ-ATP, with black and blue traces showing group averaged spectra, and red traces showing segmental integral. (C) Spectral fitting of group-averaged h-PME-corrected spectrum with Gaussian lineshape model. (D) Concentrations of brain h-PME and other metabolites for the group, in reference to γ-ATP (3.0 mM).

**Table 1.** Brain metabolite concentrations (mM) measured by 31P MRS at 7T (N = 16)

| Metabolite | Summed | Individual | Ren<sup>23</sup> | Li<sup>16</sup> | Ruhm<sup>46</sup> | Dorest<sup>26</sup> | Dorest<sup>19</sup> |
| --- | --- | --- | --- | --- | --- | --- | --- |
|  | (Gaussian) |  | 7T(Voigt) | 7T | 9.4T(WM/GM) | 9.4T | 9.4T |
| h-PME | 1.48 | 1.36±0.39 |  |  |  |  |  |
| PE | 1.64 | 1.41±0.23 | 2.27 | 1.77 | 1.94/2.56 | 0.85 | 1.40 |
| PC | 0.31 | 0.31±0.10 | 0.30 | 0.57 | 0.38/0.61 | 0.39 | 0.60 |
| Pi(ex) | 0.47 | 0.45±0.13 | 0.30 |  | 0.03/0.05 | 0.06 |  |
| Pi(in) | 1.11 | 1.05±0.15 | 0.85 | 0.99 | 1.14/1.19 | 0.70 | 1.40 |
| GPE | 0.99 | 0.88±0.14 | 0.80 | 0.57 | 1.14/1.00 | 0.66 | 1.00 |
| GPC | 1.28 | 1.12±0.15 | 1.32 | 0.84 | 1.93/1.35 | 0.99 | 1.40 |
| PCr | 4.21 | 4.24±0.50 | 4.37 | 3.66 | 3.02/3.54 | 5.07 | 4.70 |
| γ-ATP | 3.00 | 3.00 | 3.00 | 3.00 | 3.00 | 3.00 | 3.00 |
| α-ATP | 2.76 | 2.71±0.48 | 3.09 |  | 2.54/2.67 | 3.19 | 2.70 |
| NAD(H) | 0.61 | 0.53±0.21 | 0.28 | 0.57 | 0.31/0.36 | 0.76 | 0.70 |
| UDP(G) | 0.20 | 0.23±0.11 | 0.08 |  | 0.08/0.06 |  |  |
| β-ATP | 2.54 | 2.73±0.43 | 2.82 |  |  |  |  |
| PCr/(PC+GPC) | 2.64 | 2.97±0.40 | 2.70 | 2.60 | 1.31/1.81 | 3.67 | 2.35 |

Note: Data acquired using pulse acquire sequence at TR = 30 s without proton-decoupling and NOE enhancement; γ-ATP used as concentration reference (3.0 mM).
Refs (16) and (23): TR = 30 s and pulse-acquire; Ref (19): TR = 25 s and ISIS sequence; Ref (26): TR = 5 s, TM = 5 ms and STEAM sequence, with T1 and T2 correction; Ref (46): TR = 0.25 s and 3D CSI, with T1 correction.

## 4. Discussion

### 4.1 Major findings

For decades, it has been commonly thought that the brain PME 31P signals are composed of PE and PC. This study demonstrated inadequacy of such a two-component model, by revealing hidden PME signals under PE and PC resonance using Gaussian lineshape analysis and IR modulation approach. h-PME signals amount to 44 ± 9% of total PME signals in fully relaxed 31P spectra. Correction on h-PME effect is important for accurate quantification of PE and PC. The findings have implication on using PE and PC as biomarkers of altered MPL metabolism in association with brain pathologies.

### 4.2. h-PME assignment

Although it is not fully clear what metabolites contribute to the h-PME signals in the downfield PME region, the observed signal features including the integral, profile and relaxation time appear to support the hypothesis that 2,3-DPG may be a significant contributor. In human brain, the circulating blood occupies 5-6% of the intracranial space,^39, 40^ and 2,3-DPG is the most abundant in RBC at a concentration of 5-8 mM.^41, 42^ Thus, the estimated brain 2,3-DPG phosphate concentration is 8 x 0.06 x 0.5 x 2 = 0.48 mM, considering the hematocrit 50% and each 2,3-DPG possessing two phosphoryl groups. This is about half of the concentration of the measured fast-relaxing h-PME 0.98 mM (= 1.4 mM x 70%, with correction for the long-T1 component). A higher measured h-PME level might be explained by a higher vascular space in posterior brain (containing sagittal and transverse sinuses) close to the sensitivity area of the RF coil. However, unless 2,3-DPG is responsible for all h-PME signals, which means a high regional cerebral blood volume of 12%, a presence of previously unrecognized metabolites in this PME region has to be considered (see later discussion).

In terms of chemical shifts and linewidths, previous RBC 31P NMR studies indicate that the two resonances of 2,3-DPG are sensitive to environmental factors such as pH and oxygenation.^36, 37^ In fresh oxygenated blood, RBC 2,3-DPG P2 resonates at ∼5.3 ppm near Pi(ex) and P3 at 6.1 ppm near PC (Δδ = 0.83 ppm), whereas in deoxygenated blood, these two resonances are broadened and less separated (Δδ = 0.57 ppm) with P3 shifted to downfield at 6.9 ppm (near PE) and P2 at 6.3 ppm (near PC).^37^ However, under in vivo condition, 2,3-DPG signal dispersion may occur due to variations in oxygenation and/or pH.^31^ The wide distribution of the spatial orientation of blood vessels may further disperse 2,3-DPG signals via anisotropic magnetic susceptibility mechanism, an effect often manifested as an asymmetric signal slanting toward upfield^38, 43^ as seen here for h-PME in IR spectra (Figures 6 and 8).

A tentative assignment of 2,3-DPG is consistent with recent T1 and T2 findings that PC, which appears to be heavily contaminated by h-PME (Figures 3 - 7), has much shorter apparent T1 and T2 relaxation times than other MPL metabolites (PE, GPE and GPC). An examination of literature data show that the T2 shortening averaged 38% at 7T by van der Kemp et al,^31^ 28% at 9.4T by Dorst et al,^26^ and the T1 shortening averaged 32% at 7T by Ren et al.^23^ These deviations agree reasonably well with the observation of reduced PC resolution by h-PME in this study (1 - Δh/h = 0.30 – 0.55 in Figures 3-7 and 9). Confirmative evidence supports the short T1 nature of h-PME in line with RBC 2,3-DPG (T1 ∼ 2.2 s vs ∼7-9 s in solution^36^) was the observation of more rapid recovery of h-PME than PE and PC after inversion (Figures 6 and 7). Additional evidence was the observation of an elevated h-PME at long vs short TR (Figure 5), and such a TR effect is also identifiable in the brain 31P spectra recently reported by Li et al.^16^

Over the last three decades, a few attempts have been made to include 2,3-DPG in PME fittings.^32–34^ Assuming that 2,3-DPG was responsible for an additional signal between Pi and PC in some spectra collected from parieto-occipital cortex, Potwarka et al fit it by two equal-sized peaks (at 5.16 and 5.64 ppm) at fixed linewidth linked to PCr (with a +6 Hz offset).^32^ Later work by Jensen et al from the same group introduced a second metabolite (phosphoserine, resonating between PE and PC) at a fixed magnitude to stabilize and improve the fitting precision surrounding PE and PC.^33^ Very recently, Korzowski et al modeled 2,3-DPG as pseudo-doublet at a fixed separation of 0.4 ppm and allowing 2-P resonance to vary between 6.0 - 6.3 ppm with PC and PE linewidths linked to GPE and GPC.^34^ Though these efforts widely disagree in details on fitting model, they all assume that 2,3-DPG is a contributor to brain PME signals, which appears to be supported by the findings in this study, especially the observation of fast-relaxing h-PME (Figures 5-7).

However, additional or alternative sources cannot be excluded. Phosphoproteins formed by protein phosphorylation in cytosol and/or mobile phospholipids PA on cell membrane both may contribute signals in PME region, similar to mobile MPLs and lipoproteins contribution to the background signal at ∼2.2 ppm (Figure 9 and ref(16)). Especially, protein phosphorylation is the most common post-translational modification in vivo with occurrence frequency > 3-fold higher than other forms of modification combined, including acetylation, glycosylation, amidation, hydroxylation, methylation, carboxylation, ubiquitylation and sulfation.^44^ On other hand, PE and PC molecules bound to cellular macromolecules may be in slow exchange with cytosol components when shifted to a difference resonance frequencies with altered relaxation properties. Similarly, PE and PC residing in different cellular compartments with distinct pHs may also contribute signals to h-PME. Together, these sources, if present, may complicate h-PME composition.

### 4.3 Heterogeneity of h-PME profile

In addition to the major fast-relaxing h-PME signals, there is also a long-T1 component at ∼ 6.9 ppm as revealed in the subtraction spectra by the appearance of a background signal in baseline (Figure 7), which accounts for approximately 30% of the total h-PME signals. Though the assignment of this long-T1 h-PME component is as yet unclear, in vitro 31P spectra from tissue extracts suggest that G3P, a product of GPE and GPC breakdown (Figure 1), and sugar-phosphates in glycolysis pathway are potential contributors.^45^ Nevertheless, given the complexity and heterogeneity of the h-PME signals, further investigations are needed to clarify these assignments and advance our understanding of these findings.

### 4.3. h-PME effect on analysis of MPL metabolites

For major well-resolved metabolites, the current work based on Gaussian lineshape analysis yielded concentrations comparable to previous work based on Voigt lineshape mode (Table 1). However, a large difference was noticed in PE concentration (1.64 mM with h-PME correction vs 2.27 mM without correction), as expected from a large presence of h-PME (43.6 ± 8.8 % of total PME) and the fact that the chemical shift of PE (6.8 ppm) is nearly at the maximal height of h-PME profile (∼ 6.9 ppm). This means that previous Voigt lineshape fitting without adequate restriction on Lorentzian-to-Gaussian ratio among different MPL metabolites may have overestimated PE from neglecting h-PME.^23^

The PC concentration from the present study (0.31 ± 0.10 mM) is lower than those measured by localized techniques with short TR and by similar method under comparable condition but without the correction of h-PME (Table 1).^16, 46^ The [PCr]/([PC] + [GPC]) ratio from the present study (3.0 ± 0.4) is comparable to a recent measurement (2.6 ± 0.3) in healthy subjects under the similar acquisition conditions (TR = 30 s at 7T) by Li et al.^16^ This ratio was proposed to be an approximate internal standard to check 31P spectral fitting of data acquired from healthy volunteers, given the concerning variation of over an order of magnitude in the reported phosphoester levels in literature (see ref(16) and references therein). Based on Li’s formula [PCr]/([PC] + [GPC]) = 0.53 x [tCr] and the brain tCr concentration of 5.4 mM measured using 1H MRS by Zoelch et al,^47^ the expected [PCr]/([PC] + [GPC]) ratio is 2.9, which is consistent with the result from the present study. In addition to h-PME, there are a number of other factors that may cause variations in reported phosphoester levels including but not limited to cohort metabolism, pule sequence, post-processing conditions (adipozation and baseline correction), and lineshape fitting models, as noted by Ruhm et al.^46^

### 4.4. Other findings

In addition to h-PME, this study also observed two resolved Pi(ex) signals in the downfield region of brain 31P spectra, confirming our previous finding of complex composition of extracellular inorganic phosphates.^24^ The IR spectra showing h-PME profile also revealed a similar broad bump (marked as unspecified macromolecules MM, Figure 8) in the upfield of PDE region. The relative large magnitude of MM over GPE and GPC in short TI spectrum (Figure 8) indicates its short-T1 nature compared to GPE and GPC.

### 4.5 Limitations and prospects

Though the current study presented an IR-based method to filter out h-PME from reference spectra acquired by pulse-acquire sequence, there may be other alternative ways to achieve full resolution between PE and PC. For example, the short-T2 nature of 2,3-DPG can be exploited to attenuate its presence in PME by using long-echo localized pulse sequence especially at higher field,^46^ though likely at high cost of SNR and signal distortion from 1H J-modulation. It is also possible to utilize IR sequence by nulling short-T1 2,3-DPG signals with a shorter inversion delay than the one used in this study. Of course this requires an additional post-processing inversion step to bring up other inverted signals with long-T1, and a reference spectrum is still needs to calibrate the IR signals.

Finally, this study has its limitation. Due to the use of long TR (30 s) for quantitative measurements to avoid T1 correction, the data acquisition sequence was not optimal for SNR, which may in turn affect measurement precision. Acquisition at shorter TR is more practical, as research interests are increasingly shifted toward 3D MRSI for its high spatial resolution in resolving metabolic abnormalities pertain to pathology. Therefore, further study is needed to develop method optimized for short TR and extend it to the other platforms specially 3T, which has a much larger installation base than 7T. On the other hand, the new finding of the h-PME signals may permit us to develop and test new hypotheses, and to obtain new information about metabolism of challenging brain diseases such as cancers, stroke and Alzheimer’s diseases.

## 5. Conclusion

Using lineshape analysis and inversion-recovery modulation, this study demonstrated the presence of hidden PME signals under PE and PC resonances in brain 31P MR spectra in vivo. Fast relaxing h-PME signals is tentatively assigned to RBC 2,3-DPG, though other sources of contribution cannot be excluded. h-PME correction is needed for accurate quantification of PE and PC and using them as biomarkers of altered phospholipid metabolism in brain pathologies.

## Data Availability

All data produced in the present study are available upon reasonable request to the authors

## Acknowledgements

The authors are grateful for helpful discussion with Dr. A. Dean Sherry, 7 T operational assistance from Corey Mozingo and Pena Sal, and technical support from Dr. Ivan Dimitrov (Philips Healthcare).

