## Supplementary Material Table 1S for "31P-MRS of healthy human brain: revealing the hidden PME signals under phosphoethanolamine and phosphocholine resonances at 7T"

Table 1S. Linewidth of brain metabolite 31P signals measured at 7T

| **Metabolite** | **Summed** | **Individual** | **Ren et al** | **Dorst et al** |
| --- | --- | --- | --- | --- |
|  |  | Mean ± std | Ref(23) | Ref(47) |
| h-PME | 105.0 | 105.3 ± 25.4 |  |  |
| PE | 21.5 | 21.3 ± 2.0 | 28.9 | 21.3 |
| PC | 19.5 | 23.52 ± 8.4 | 25.5 | 24.1 |
| Pi(ex) | 43.9 | 37.2 ± 14.1 | 39.5 |  |
| Pi(in) | 25.4 | 25.6 ± 2.9 | 26.9 | 19.9 |
| GPE | 22.5 | 21.6 ± 3.2 | 24.4 | 20.9 |
| GPC | 19.5 | 21.9 ± 2.5 | 25.5 | 20.6 |
| PCr | 13.7 | 13.6 ± 1.6 | 19.7 | 9.8 |
| γ-ATP | 49.8 | 46.0 ± 6.2 | 49.8 | 44.0 |
| α-ATP | 37.1 | 36.5 ± 0.5 | 41.2 | 40.7 |
| NAD(H) | 51.8 | 47.9 ± 16.4 | 51.7 |  |
| UDP(G) | 50.8 | 35.9 ± 12.2 | 41.3 |  |
| β-ATP | 66.4 | 74.5 ± 11.8 | 74.2 |  |

Note: Data acquired by pulse-acquire sequence at TR = 30 s. Ref(23) data corrected with a ratio factor (γ_31P_/γ_1H_). Ref(47) data acquired at 9.4T by STEAM sequence with TR = 5 s.
